# The role of IgM testing in the diagnosis and post-treatment follow-up of syphilis: a prospective cohort study

**DOI:** 10.1101/19011148

**Authors:** Kara K. Osbak, Achilleas Tsoumanis, Irith De Baetselier, Marjan Van Esbroek, Hilde Smet, Chris R. Kenyon, Tania Crucitti

**Author notes:** these authors contributed equally to this work (Kara K. Osbak, Achilleas Tsoumanis, Chris R. Kenyon, Tania Crucitti). **Corresponding author:** KKO: Nationalestraat 155, 2000, Antwerp, Belgium.

## Abstract

**Objectives:** The diagnosis of repeat syphilis and its follow-up remains challenging. We aimed to investigate if IgM testing may assist in the diagnosis of syphilis reinfection/relapse and its treatment follow-up.

**Methods:** Sera were collected from 120 individuals with a new diagnosis of syphilis (72 with repeat infections) and 30 syphilis negative controls during a cohort study investigating syphilis biomarkers conducted at a STI/HIV clinic in Antwerp, Belgium. Syphilis was diagnosed based on a simultaneous positive treponemal and non-treponemal assay result and/or positive serum PCR targeting *polA*. Specimens collected at visit of diagnosis, and 3 and 6 months post-treatment were tested by two enzyme immuno assays (EIAs), recomWell (Mikrogen; MI) and Euroimmun (EU), to detect anti-treponemal IgM. Baseline specimens were also tested for anti-treponemal IgM using a line immuno assay (LIA) recomLine (Mikrogen). Quantitative kinetic decay curves were constructed from the longitudinal quantitative EIA results.

**Results:** An overall sensitivity for the diagnosis of syphilis of 59.8 % (95 % confidence interval (CI): 50.3-68.7), 75.0 % (95 %(CI): 66.1-82.3) and 64.2 % (95 %(CI): 54.8-72.6) was obtained for the EU, MI EIAs, and MI LIA, respectively. When only considering repeat syphilis, the diagnostic sensitivity decreased to 45.7 % (95 %(CI): 33.9-58.0), 63.9 % (95 %(CI): 51.7-74.6) and 47.2 % (95 %(CI): 35.5-59.3), respectively. IgM seroreverted in most cases 6 months after treatment. Post-treatment IgM concentrations decreased 30 % faster for initial syphilis compared to repeat infection. The IgM EIAs and IgM LIA agreed from fairly to moderately (Cohen’s kappa (κ): 0.36 [EU EIA]; κ: 0.53 [MI EIA]; κ: 0.40 [MI LIA]) with the diagnosis of syphilis.

**Conclusions:** IgM detection was not a sensitive method to diagnose syphilis and was even poorer in the diagnosis of syphilis repeat infections, but might play a role in treatment follow-up.

**KEY MESSAGE BOX:**

- IgM detection was not a sensitive method to diagnose syphilis and was even poorer in the diagnosis of syphilis repeat infections
- IgM serorevert in most cases 6 months after treatment
- In repeat episodes of syphilis, IgM concentrations decline faster post-treatment

## INTRODUCTION

Syphilis has reemerged during the last 15 years as a major public health problem with increasing incidence particularly amongst men who have sex with men (MSM) in the northern hemisphere. [1] It is a multistage chronic disease caused by *Treponema pallidum* subspecies *pallidum*, and can be challenging to diagnose, especially very early and repeat infections. Diagnostic strategies remain reliant on serological testing, however, deficiencies in assay performance and persistent anti-treponemal antibody presence following initial infection often hamper diagnostic accuracy. [2]

As the epidemics progress, an increasing proportion of all infections have been noted to be reinfections. [3] Since reinfections are more likely to present asymptomatically [4,5], timely diagnosis depends on the diagnostic accuracy of serological tests.

Since treponemal tests (TT) remain positive for life, the diagnosis of reinfections typically depends on fourfold or greater rises in non-treponemal test (NTT) titers, such as the Rapid Plasma Reagin (RPR) test. Conversely, a fourfold decline in NTT titers is used to determine the success of syphilis treatment. Due to the aspecific nature of NTTs, that relies on antibody binding to lipoidal components released during host cell destruction and also present in low quantities in the *T. pallidum* cell wall [6], other biological, infectious or immunological mechanisms can also result in fluctuations in NTT leading to false positive results. [7]

Following an initial infection, immunoglobulin (Ig) M antibodies are the first class of antibodies produced. Early detection of IgM could therefore help with the diagnosis of early syphilis. Current European syphilis guidelines [8] mention that IgM testing is useful in the assessment of newborns and cerebral spinal fluid. These guidelines also note that a negative IgM result cannot exclude the diagnosis of congenital or neurosyphilis. A study published in 2013 [9] evaluated three commercially available IgM enzyme immune assays (EIAs) using 307 serum samples from individuals with active syphilis. It found that these IgM assays had a median sensitivity of 84.5 % with specificities in the range of 91.4 - 100 %. In 23/59 (39 %) of cases the IgM EIA test was positive in suspected very early infection where the NTT was negative. A recently published prospective analysis of IgM among a cohort of MSM demonstrated generally low EIA IgM test performance for repeat syphilis diagnosis, namely only 38.5 % of repeat syphilis cases were diagnosed correctly with IgM testing [10]. The number of patients with repeat syphilis in the study was only 13.

In this study we investigated the clinical utility of IgM detection in the diagnosis of syphilis and post-treatment follow-up in a prospective cohort of 120 individuals with a new episode of syphilis. In addition we evaluated the performance of three commercially i.e. two EIAs and one Line Immmuno Assay (LIA), IgM assays.

## MATERIALS AND METHODS

### Study design

This sub-study was conducted in the context of a syphilis biomarker discovery study (ClinicalTrials.gov Nr: NCT02059525). Potentially eligible study participants 18 years or older, in whom a new syphilis diagnosis was made, were consecutively screened and prospectively recruited between January 2014 and August 2015 at a sexually transmitted infection/ HIV clinic in Antwerp, Belgium. Study exclusion criteria were the use of beta-lactam, doxycycline or macrolide antibiotics during the 28 days preceding enrollment. Syphilis diagnosis and disease staging was performed according to the Centers for Disease Control guidelines. [11] Reinfection was defined as previously described. [3] Stage-appropriate treatment was administered according to European guidelines. [12] All participants with syphilis were followed-up at 3 and 6 months post-treatment. The study physician recorded clinical details and laboratory results in a standardized fashion during each study visit. HIV-infected controls with both negative NTT and TT results were included during the same recruitment period.

### Clinical serological testing during routine workout

Blood was drawn into serum gel tubes (Sarstedt Monovette, Nümbrecht, Germany). Sera were divided and either 1) stored at 4-8 °C until routine syphilis serological testing within four days or 2) stored at −80 °C within three hours for later testing. Routine serological testing included Macro-Vue RPR Card (RPR) (Becton Dickinson, Sparks, MD, United States (US)) and TPA assay (Ortho-Clinical Diagnostics, Rochester, NY, US) testing following the manufacturer’s instructions. Positive RPR results were determined to a titer endpoint. Sera obtained at baseline were also tested with the SERODIA-TPPA (Fujirebio Inc., Tokyo, Japan) assay and an *in-house T. pallidum* PCR targeting *polA*. [13]

### EIA and LIA testing for IgM serum antibodies

In order to investigate if assays from different manufacturers or using another method performed differently and thus would have an impact on our study outcome, we evaluated side by side two IgM EIAs and one LIA.

The anti-*Treponema pallidum* IgM EIAs were provided by Euroimmun (Lübeck, Germany) and by Mikrogen GmbH (Neuried, Germany), henceforth referred to as ‘EU EIA’ and ‘MI EIA’, respectively. Both assays use microplate wells coated with a mixture of four antigens of *T. pallidum*: Tp15, Tp17, Tp47 and TmpA. Testing was performed following the manufacturer’s instructions (EI_2111M_A_UK_C07.doc; version: 08/09/2011 and GIRETP011DE.doc; version April 2010). All quantitative EIA results were expressed in International Units (IU)/mL.

The recomLine Treponema line immune assay (LIA) kit (MI LIA) (Mikrogen GmbH) was used for the qualitative determination of the IgM antibodies for baseline samples. This test uses recombinant *T. pallidum* antigens fixed on nitrocellulose membrane strips. Testing was performed according to the manufacturer’s instructions (GARLTP002EN, version 2012/08). Objective reading of the strips was done by scanning the strips with a flatbed scanner and analysis software Recomscan (Mikrogen). The density of the antigen was compared to a positive control and ratio’s were calculated. The assay was reported negative of no antigen was detected (ratio <1), was borderline only one random antigen was detected (ratio ≥ 1) and positive if at least two random antigens were present (ratio ≥ 1).

### Quality control

The samples were analyzed by a research and diagnostic laboratory unit, both are ISO15189 accredited. One single lotnumber was used for all EIAs and LIAs. Two laboratory technicians performed the analyses during two batch testing periods of 14 days in 2016. Evaluative testing of the EIAs took place maximum 974 days after the samples of interest were collected. Samples were thawed, tested and refrozen on the same day with a maximum of two freeze thaw cycles.

Due to the prolonged testing period and nature of the routine diagnostic laboratory setting, the reagents used for the TPPA, TPA and RPR tests were from different lot numbers.

The laboratory technicians were blinded from the patient’s clinical information and any other syphilis serology result.

Data was manually double entered into the database.

### Statistical analysis

In the absence of a single gold standard test for syphilis, a positive RPR test, or in the case of reinfection a four-fold increase in RPR titer, together with a positive TPPA/TPA results or a positive *T. pallidum* PCR test were used to define a new syphilis episode (‘syphilis diagnosis’).

Percentage agreement and Cohen’s kappa (κ)-coefficient value [14] were calculated to estimate agreement between the IgM test results. The sensitivity, specificity, positive (PPV) and negative (NPV) predictive values with 95 % confidence intervals (CIs) were calculated using the baseline visit samples. Borderline results for the EIA and LIA testing were included for the first set of analyses as negative (specific scenario) and the second dataset included all borderline results as positive (sensitive scenario).

Continuous variables were expressed as median values and interquartile range (IQR). Associations between categorical variables were assessed with the chi-squared test (χ^2^) and Fisher’s exact test for small numbers. A student t-test was performed to compare the quantitative results of the IgM assays between initial and repeat syphilis groups. Analyses were performed in Stata 13.1 (StataCorp LP, College Station, TX, US). The statistical significance level was set at 0.05.

Non-linear models were used to visually assess the decay of the IgM concentrations and RPR titers over time. The exponential decay curves were of the form Y = a*e^-b*X^, where Y was the measurement of the EIA tests and X the time in months. The exponential decay curves and the graphics were designed using R version 3.4.4. [15]

## RESULTS

### Study participants

In total 150 individuals were included in the study, 120 diagnosed with syphilis and 30 controls. [16] Study subject characteristics are described in **Table 1**. Of those who had active syphilis at the time of study enrollment 48/120 (40 %) presented with a first episode of syphilis (henceforth referred to as ‘initial infection’) and 72/120 (60 %) presented with a repeat infection. Previous NTT results were available for 107/120, the remaining 13 individuals without previous serological test results were classified as having an initial infection based on the fact that they had never had a diagnosis of or symptoms suggestive of syphilis before. Initial infections were more often symptomatic 34/48 (71 %) compared to the repeat infections 39/72 (54 %) (P = 0.09). [5]

**Table 1.** Study subject characteristics

| | **Syphilis-**<br>**positive**<br>**cases**<br>**N=120** | **Controls<sup>\$</sup>**<br>**N=30** |
| --- | --- | --- |
| **Characteristics** | **n (%) / median**<br>**(IQR)** | **n (%) / median**<br>**(IQR)** |
| **Gender (male)** | 119 (99) | 30 (100) |
| **MSM** | 117 (98) | 24 (80)** |
| **HIV-infected** | 103 (86) | 30 (100)** |
| **Taking ART** | 91 (88) | 24 (80) |
| **Benzathine**<br>**Penicillin G**<br>**treatment** | 117 (98) | NA |
| **RPR-C titer at**<br>**baseline** | 64 (32-128) | 0 |
| **Age** | 40 (31.5-48) | 37.5 (32-45) |
| **CD4<sup>+</sup> T-cell count** | 598 (448-749) | 577 (392-684) |
| ***Syphilis Stages and Infection History*** |  |  |
| **Syphilis Stage** | **Initial**<br>**Infection \*** | **Repeat**<br>**Infection \*** |
| **Primary** | 12 (10) | 11 (9) |
| **Secondary** | 22 (18) | 28 (23) |
| **Early Latent** | 7 (6) | 24 (20) |
| **Late Latent** | 7 (6) | 9 (8) |

*Legend: ART- antiretroviral therapy; NA- not applicable; \*- percentage calculated with denominator N = 120 syphilis positive subjects; \$- control subjects were all HIV positive and non-treponemal and treponemal antibody negative at the time of study inclusion; \*\*- statistically significant difference between syphilis positive group and controls*

### IgM test results

The IgM testing was performed on 339, 343 and 150 sera by the EU, MI EIAs and LIA, respectively. Details of the qualitative results are presented in **Table 2**. One sample from a control patient tested IgM positive and another one tested borderline positive with the MI EIA. The LIA also had one false positive and one borderline result. All control samples tested negative with the EU EIA.

**Table 2.** Qualitative enzyme immunoassay and line immunoassay results

| Characteristic | EU EIA |  |  | MI EIA |  |  | MI LIA |  |  |
| --- | --- | --- | --- | --- | --- | --- | --- | --- | --- |
|  | Total | Positive | Borderline | Total | Positive | Borderline | Total | Positive | Borderline |
|  | N | N (%) | N (%) | N | N (%) | N (%) | N | N (%) | N (%) |
| ***Time of testing*** |  |  |  |  |  |  |  |  |  |
| **Baseline** | 147 | 70 (48) | 9 (6) | 150 | 91 (61) | 4 (3) | 150 | 77 (51) | 19 (13) |
| **3M** | 86 | 16 (19) | 10 (12) | 86 | 37 (43) | 1 (3) | NT | NT | NT |
| **6M** | 106 | 16 (15) | 6 (6) | 107 | 24 (22) | 15 (14) | NT | NT | NT |
| ***Syphilis Stage/ Controls at baseline visit*** |  |  |  |  |  |  |  |  |  |
| **Primary** | 23 | 14 (61) | 3 (13) | 23 | 19 (83) | 0 (0) | 23 | 17 (74) | 2 (9) |
| **Primary (repeat only)** | 11 | 5 (46) | 1 (9) | 11 | 7 (64) | 0 | 11 | 7 (64) | 0 |
| **Secondary** | 48 | 33 (69) | 3 (6) | 50 | 43 (86) | 0 (0) | 50 | 38 (76) | 6 (12) |
| **Secondary (repeat only)** | 27 | 13 (48) | 2 (7) | 28 | 21 (75) | 0 | 28 | 16 (75) | 6 (21) |
| **Early Latent** | 30 | 14 (47) | 1 (3) | 31 | 16 (52) | 3 (10) | 31 | 11 (35) | 8 (26) |
| **Early Latent (repeat only)** | 23 | 10 (43) | 1 (4) | 24 | 11 (46) | 3 (13) | 24 | 6 (25) | 8 (33) |
| **Late Latent** | 16 | 9 (56) | 2 (13) | 16 | 12 (75) | 0 (0) | 16 | 10 (63) | 2 (13) |
| **Late Latent (repeat only)** | 9 | 4 (44) | 2 (22) | 9 | 7 (78) | 0 | 9 | 5 (56) | 2 (22) |
| **Controls** | 30 | 0 (0) | 0 (0) | 30 | 1 (3) | 1 (3) | 30 | 1 (3) | 1 (3) |
| ***Syphilis History*** |  |  |  |  |  |  |  |  |  |
| **Initial Infection** | 47 | 38 (81) | 4 (9) | 48 | 44 (92) | 0 (0) | 48 | 42 (88) | 2 (4) |
| **Repeat Episode** | 70 | 32 (46) | 5 (7) | 72 | 46 (64) | 3 (4) | 72 | 34 (47) | 16 (22) |

Legend: NT- not tested. EU: Euroimmun; MI: Mikrogen; LIA: line immunoassay; EIA: enzyme immunoassay
- Note: 1. the number of baseline samples includes the 30 samples of the control syphilis negative individuals 2. the number of positive MI EIA and MI LIA results includes 1 positive result obtained among the control samples 3. the number of borderline MI EIA and MI LIA results includes 1 borderline result obtained among the control samples

### IgM test performs suboptimally for diagnosis of syphilis

The overall diagnostic performance for the two IgM EIAs and LIA assays evaluated was moderate. **Table 3** summarizes the assay performances in the specific case scenario. The diagnostic sensitivities of the evaluated assays ranged from 80.9 % to 91.7 % when applied to samples collected at baseline from individuals with initial syphilis. When baseline analyses were stratified per initial or repeat infection, the diagnostic sensitivity decreased significantly in repeat infections for all assays evaluated compared to initial infection (P=0.002 (MI EIA); P=0.003 (EU EIA);P<0.0001 LIA MI).

**Table 3.**
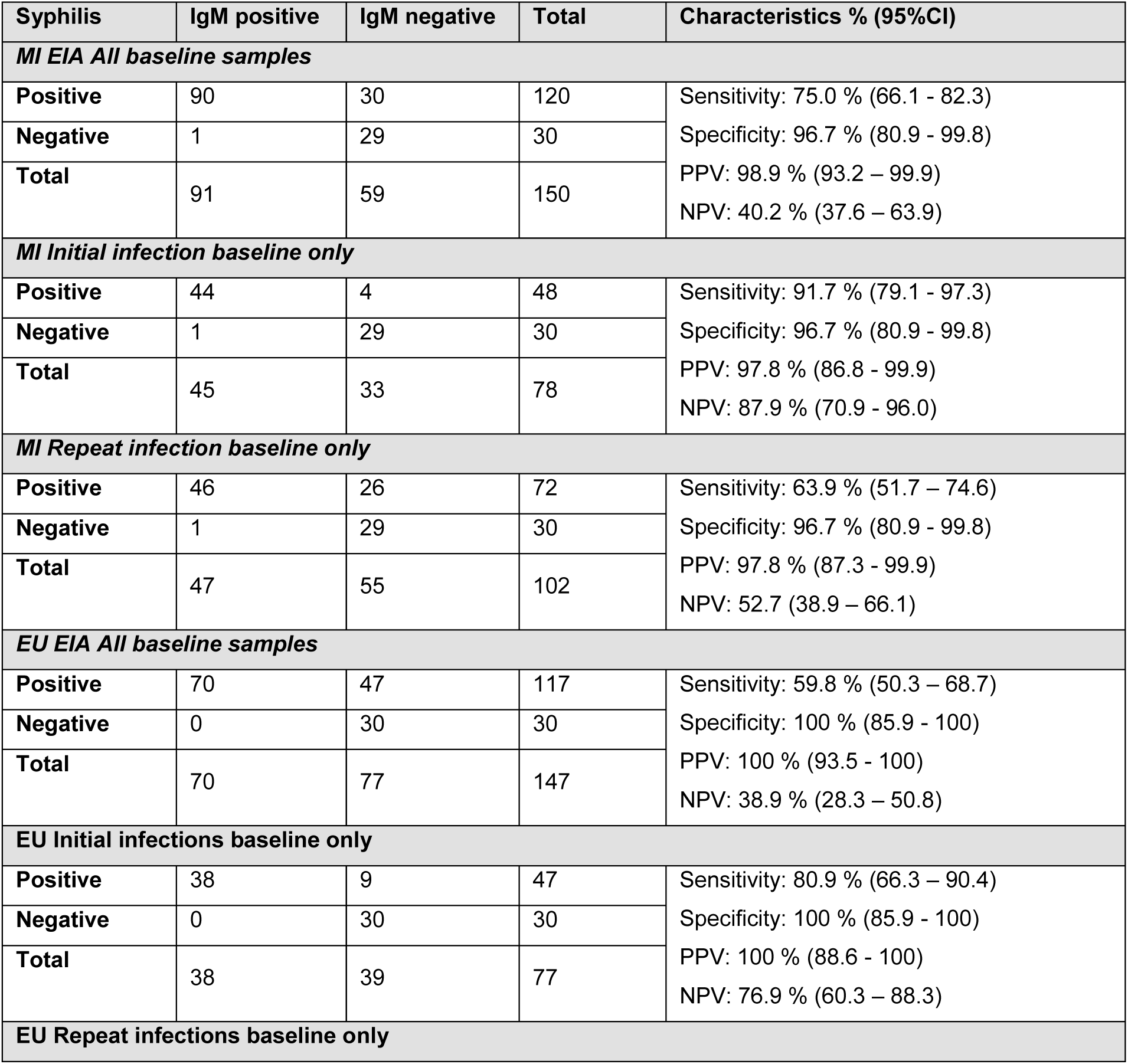
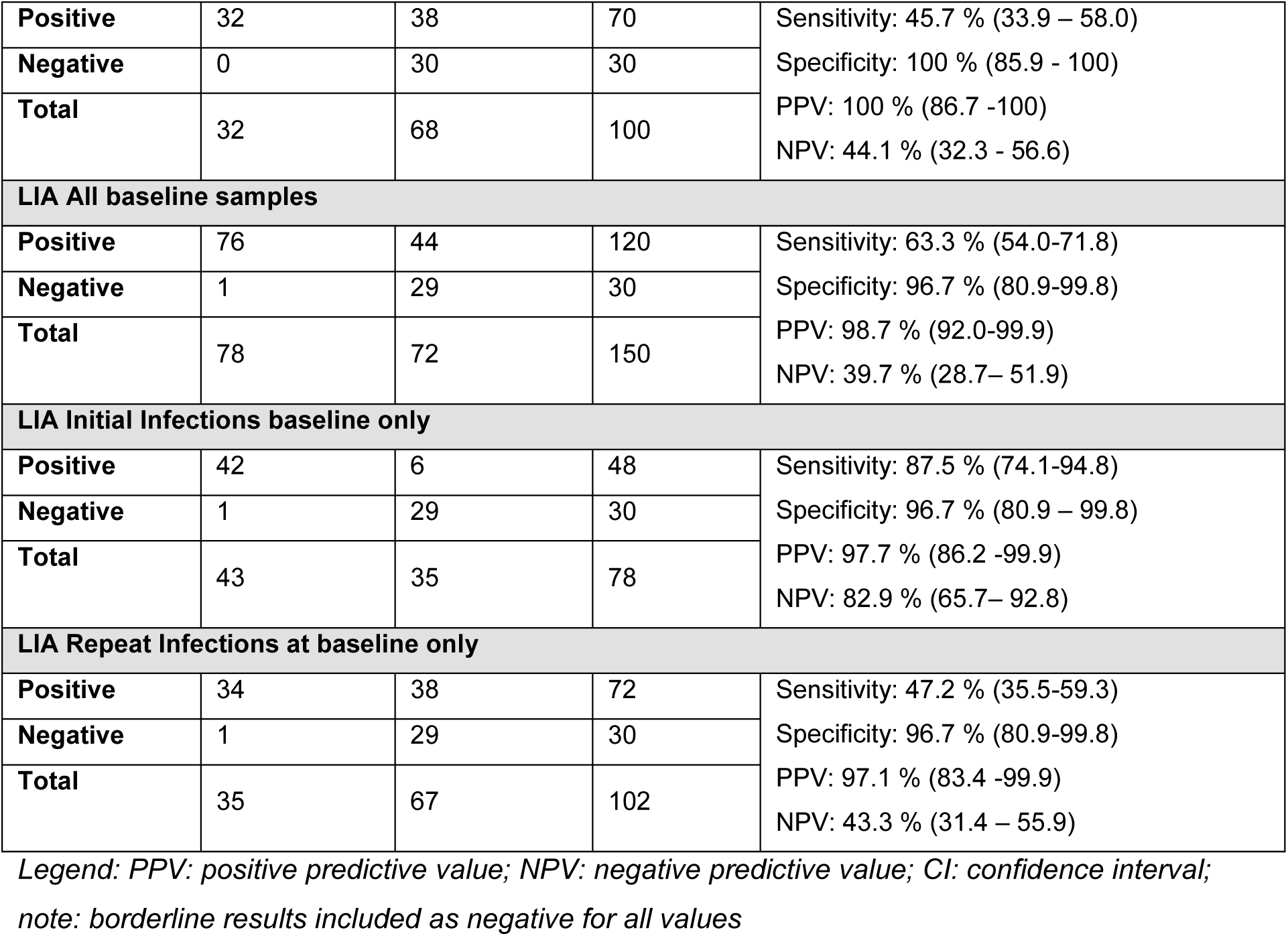
Cross-tabulations and performance characteristics of the serological assays (EIAs/LIA) tested in this study according to the specific scenario*

The performances of the assay in the sensitive-case scenario are presented in Supplementary table S1

### LIA IgM testing of baseline samples

The overall performance of the LIA test was moderate (Table 2 & 3). Of the six antigen lines evaluated, Tp47 and TmpA were the most frequently positive. No samples were positive for the marker Tp257. A heat map was created comparing the RPR titer and MI LIA (Supplementary figure S2).

### Quantitative assessment of baseline samples with EIA

When considering the quantitative results of the two EIAs, a significant difference was found between the initial and repeat syphilis groups (EU P < 0.001; MI P = 0.001) with repeat syphilis having lower IgM concentrations. Moreover, the concentrations were significantly lower for individuals presenting with latent stage syphilis compared to primary and secondary stage (EU EIA P = 0.02; MI EIA P = 0.004).

### Longitudinal post-treatment follow-up at 3- and 6-month and kinetic decay curve characteristics of EIA testing

At 6 months post-treatment seroreversion of IgM test results occurred in 57/70 (81 %) samples tested by the EU EIA and 66/90 (73 %) tested by MI EIA. Among the month 6 samples for which a seroreversion was not observed, 6/13 and 7/24 tested with the EU EIA and MI EIA, respectively, did not show a decrease in RPR titer either. Overall, in a total of 19 individuals a decrease in RPR titer post-treatment was not observed. Individuals with an initial syphilis infection demonstrated a faster decrease in analyte concentration for every additional time unit (0.55 (95 % CI: 0.36 – 0.68)) compared to a repeat syphilis (0.74 (95 % CI: 0.62 – 0.83))(Figure 1).

**Figure 1.**
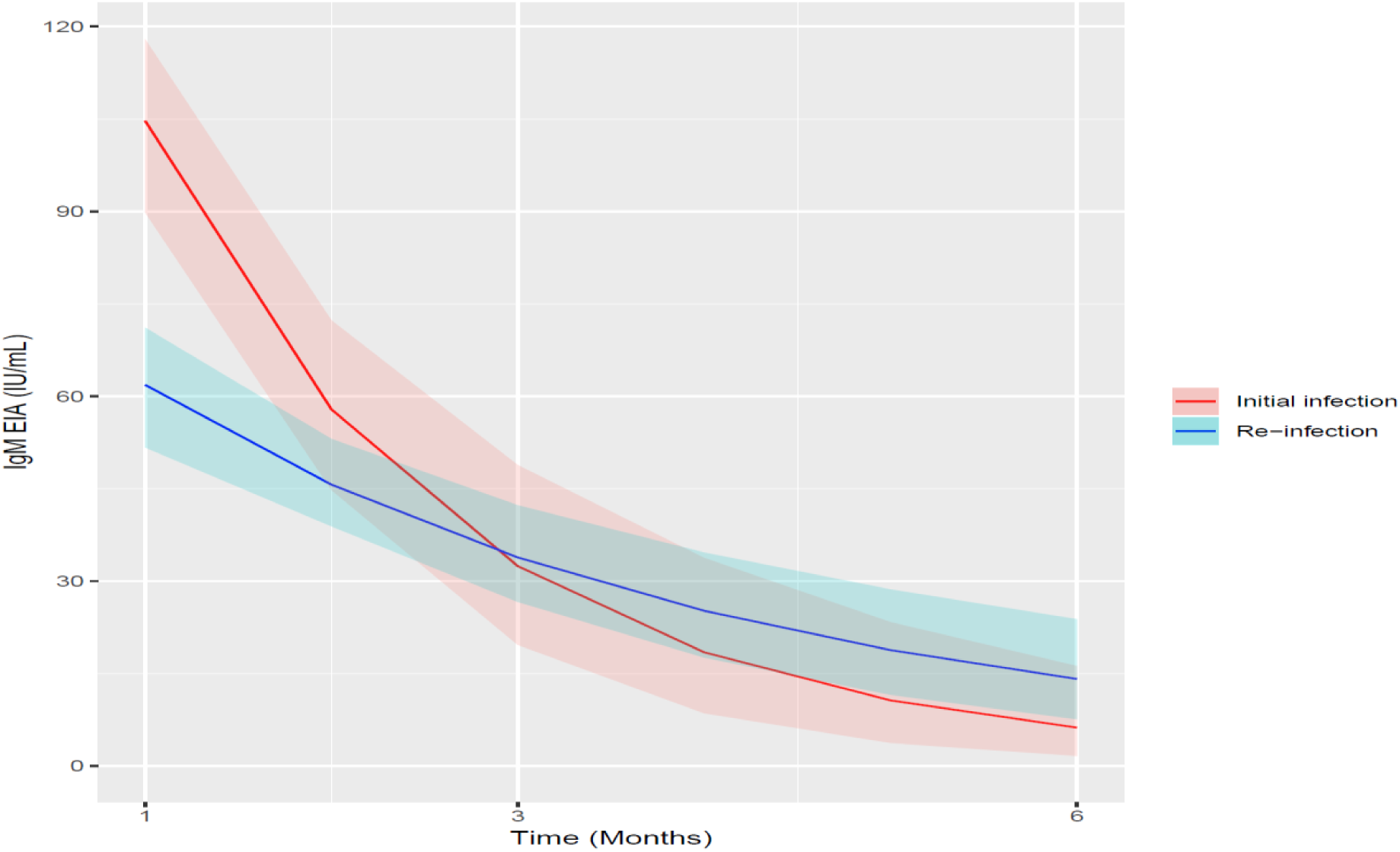
Initial and post-treatment decay curves of recomWell EIA IgM

Supplementary figure S3 represents the decay curves of IgM EIA Mikrogen concentration and RPR titers over time.

### Agreement between the commercial IgM tests

The overall agreement was substantial between the two EIA tests when considering all samples i.e. samples from baseline, M3, and M6, tested by both assays (N = 339). The Cohen’s kappa was κ: 0.69 (85.3 %) and κ: 0.74 (86.7 %) for the specific-case and sensitive-case scenarios, respectively.

There was a strong agreement between the LIA and the EIAs: 90.7 % (κ: 0.81) between the MI EIA and MI LIA, and 95.3 % (κ: 0.91) between EU EIA and MI LIA in the specific-case scenario. In the sensitive-case scenario the agreement between MI EIA and MI LIA increased further up to 99.3 % (κ: 0.91) but decreased between EU EIA and MI LIA to 88.7 % (κ: 0.77).

## DISCUSSION

We aimed to determine whether testing for IgM could aid in the diagnosis of new syphilis infections, including repeat infections, and if it could be useful for post-treatment follow-up. Detection of IgM may help in the diagnosis of syphilis but its diagnostic sensitivity is poor, it was notably low in participants diagnosed with repeat syphilis. Our results are, however, congruent with previous reports of suboptimal IgM test performance overall and for repeat syphilis in particular. [10] The lower diagnostic sensitivity of IgM in repeat syphilis is compatible with results from other infections where reinfections lead directly to increases in IgG without initial increases in IgM. [17]

This study represents the most comprehensive evaluation of EIA and LIA IgM testing on serum from individuals with syphilis that we are aware of. We found that both EIA agreed substantially, albeit that the Mikrogen EIA performed in terms of diagnostic sensitivity better than the Euroimmun assay. The difference between both sandwich EIAs lays in the pre-treatment of serum with the IgG/RF absorbant included in the EIA reagent kit of Euroimmun. The absorbant removes the antibodies of the IgG classes and the rheumatoid factors, known to be possible interferences in IgM EIA. False positive reactions may occur in pathogen specific IgM detection when the IgM rheumatoid factors bind to IgG immune complexes. On the other hand, false negative results may appear when pathogen specific IgM antibodies are displaced by stronger binding IgG. [18] We cannot rule out that this extra IgG/RF removal step may have contributed to the differences in test performance found between both EIAs.

The general performance of the LIA for the baseline syphilis diagnosis was similar to the EIAs. The antigen line reaction varied by sample and in some samples only a few antigens could be demonstrated. Since no particular single antigen or line can be attributed to a stage or profile, an overall qualitative result should be considered.

In the majority of the IgM positive cases detected at baseline the IgM antibodies disappeared 6 months after treatment. Interestingly the decline in IgM concentrations occurred faster in initital syphilis compared to repeat syphilis. Our study hints but remains inconclusive and additional studies are required to investigate whether IgM concentration measurements may be a more objective, usefull and high throughput method compared to RPR for the follow-up of syphilis treatment.

More studies are needed to investigate the role of IgM in syphilis diagnosis on a more diversified and larger scale (e.g. women, non-MSM populations). Since our study only included individuals with a dual positive NTT/TT we were unable to evaluate if IgM values might precede NTT/TT seroconversion. A previous study did find that IgM could be useful in the early diagnosis of syphilis in the subset of patients with equivocal TT and negative NTTs. [9]

Strengths of this study include the comprehensive characterization of sera and its prospective nature. Limitations to this study include the fact that the study population was mostly HIV infected. We did not control for possible effects of HIV-infection such as viral load and CD4^+^ T-cell count, although notably most HIV-infected participants were taking ART and had a high immune cell count thus the likelihood of this having an effect on the test outcome is unlikely. The study was not designed to adequately evaluate the assays’ specificity. [19]

## Conclusions

In conclusion, the clinical utility of IgM testing for syphilis diagnosis is suboptimal. Low diagnostic sensitivity, especially in the case of repeat syphilis could lead to missed infections. However, IgM might have some utility in the follow-up of treatment.

## Data Availability

The data supporting the findings of this publication are retained at the Institute of Tropical Medicine (ITM), Antwerp and will not be made openly accessible due to ethical and privacy concerns. According to the ITM research data sharing policy, only fully anonymised data can be shared publicly. The data are de-identified (using participant identification numbers only) but not fully anonymised and it is not possible to fully anonymise them due to the longitudinal nature of the data. Data can however be made available after approval of a motivated and written request to the ITM at ITM. The ITM data access committee will verify if the dataset is suitable for obtaining the study objective and assure that confidentiality and ethical requirements are in place.

## ABBREVIATIONS

ART: Antiretroviral therapy
RPR: rapid plasma reagin card test
NTT: non-treponemal test
TT: treponemal test
HIV: Human Immunodeficiency Virus
ITM: Institute Tropical Medicine-Antwerp
MSM: Men who have Sex with Men
TPPA: *Treponema pallidum* passive particle agglutination test
EIA: enzyme immunoassay
LIA: line immunoassay

## AUTHOR STATEMENTS

## Acknowledgements

We would like to thank the ITM laboratory teams for their involvement in the study and the authors gratefully acknowledge the individuals who participated in this study. Mikrogen and Euroimmune generously provided the tests used in this study. Parts of this work have been presented at IUSTI and ECCMID conferences.

## Competing Interests

The authors declare no conflicts of interest. The sponsors had no role in the design of the study; in the collection, analyses, or interpretation of data; in the writing of the manuscript, and in the decision to publish the results.

## Funding information

This work was part of Project ID: 757003 funded by the Flemish Government-Department of Economy, Science & Innovation granted to CRK. The anti-*Treponema pallidum* IgM assays were provided free of charge by Euroimmun (Lübeck, Germany) and by Mikrogen GmbH (Neuried, Germany).

## Ethical Statement

The Institutional Review Board of the ITM and the Ethics Committee of the University Hospital Antwerp approved this study (13/44/426). Written informed consent for study participation and reporting of anonymized clinical details was obtained from all participants upon study inclusion.

## Contributions

KO, IDB, MVE, CR and TC conceived the study. HS and KO co-ordinated and performed laboratory analyses and entered data. MVE, IDB and TC supervised laboratory activities. KO and AT managed and conducted the data analyses. KO wrote the first draft. All authors contributed to the final version of the manuscript and approved the final manuscript.

## Data availability

The data supporting the findings of this publication are retained at the Institute of Tropical Medicine (ITM), Antwerp and will not be made openly accessible due to ethical and privacy concerns. According to the ITM research data sharingpolicy, only fully anonymised data can be shared publicly. The data are de-identified (using participant identification numbers only) but not fully anonymised and it is not possible to fully anonymise them due to the longitudinal nature of the data. Data can however be made available after approval of a motivated and written request to the ITM at. The ITM data access committee will verify if the dataset is suitable for obtaining the study objective and assure that confidentiality and ethical requirements are in place.

## SUPPLEMENTARY INFORMATION

**S1 Table**. Cross tabulations and performance characteristics of the serological assays (EIAs/LIA) tested in this study and including borderline results as positive (sensitive-case scenario)

**S2 Figure**. Heat map comparing RPR and MI LIA results 1

**S3 Figure**. RPR/IgM decay curves of recomWell EIA IgM

## Notes

### Competing Interest Statement

The authors have declared no competing interest.

### Clinical Trial

ClinicalTrials.gov Nr: NCT02059525

## REFERENCES

1 Abara WE, Hess KL, Neblett Fanfair R, et al. Syphilis Trends among Men Who Have Sex with Men in the United States and Western Europe: A Systematic Review of Trend Studies Published between 2004 and 2015. PLoS One 2016;11:e0159309. doi:10.1371/journal.pone.0159309

2 Jost H, Castro A, Cox D, et al. A comparison of the analytical level of agreement of nine treponemal assays for syphilis and possible implications for screening algorithms. BMJ Open 2013;3:e003347. doi:10.1136/bmjopen-2013-003347

3 Kenyon C, Lynen L, Florence E, et al. Syphilis reinfections pose problems for syphilis diagnosis in Antwerp, Belgium - 1992 to 2012. Euro Surveill 2014;19:20958.

4 Kenyon C, Osbak KK, Apers L. Repeat syphilis is more likely to be asymptomatic in HIV-infected individuals: A retrospective cohort analysis with important implications for screening. Open Forum Infect Dis Published Online First: 2018. doi:10.1093/ofid/ofy096

5 Kenyon C, Tsoumanis A, Osbak K, et al. Repeat syphilis has a different immune response compared with initial syphilis: an analysis of biomarker kinetics in two cohorts. Sex Transm Infect 2017;:sextrans-2017-053312. doi:10.1136/sextrans-2017-053312

6 Borkhardt H-L, Zielinski S. Influence of cardiolipin antibodies on the binding of treponemal specific antibodies in the fluorescence treponemal antibody absorption test and the Treponema pallidum immobilisation test. J Med Microbiol 1997;46:965–72. doi:10.1099/00222615-46-11-965

7 Nandwani R, Evans DT. Are you sure it’s syphilis? A review of false positive serology. Int J STD AIDS 1995;6:241–8. doi:10.1177/095646249500600404

8 Janier M, Hegyi V, Dupin N, et al. 2014 European guideline on the management of syphilis. J Eur Acad Dermatology Venereol 2014;28:1581–93. doi:10.1111/jdv.12734

9 Bosshard PP. Usefulness of IgM-specific enzyme immunoassays for serodiagnosis of syphilis: Comparative evaluation of three different assays. J Infect 2013;67:35–42. doi:10.1016/j.jinf.2013.03.011

10 Pastuszczak M, Kotnis-Gąska A, Jakubowicz B, et al. Utility of antitreponemal IgM testing in the diagnosis of early and repeat syphilis among HIV-infected and non-infected patients. Int J STD AIDS 2018;:095646241876284. doi:10.1177/0956462418762849

11 Workowski KA, Berman S, Centers for Disease Control and Prevention (CDC). Sexually transmitted diseases treatment guidelines, 2010. MMWR Recomm reports Morb Mortal Wkly report Recomm reports 2010;59:1–110.

12 French P, Gomberg M, Janier M, et al. 2008 European Guideline on the Management of Syphilis Date: 1. World Health 2012.

13 Liu H, Rodes B, Chen C-Y, et al. New Tests for Syphilis: Rational Design of a PCR Method for Detection of Treponema pallidum in Clinical Specimens Using Unique Regions of the DNA Polymerase I Gene. J Clin Microbiol 2001;39:1941–6. doi:10.1128/JCM.39.5.1941-1946.2001

14 Cohen J. A Coefficient of Agreement for Nominal Scales. Educ Psychol Meas 1960;20:37–46. doi:10.1177/001316446002000104

15 Team RC. R: A language and environment for statistical computing. 2018.

16 Osbak K, Abdellati S, Tsoumanis A, et al. Evaluation of an automated quantitative latex immunoturbidimetric non-treponemal assay for diagnosis and follow-up of syphilis: a prospective cohort study. J Med Microbiol 2017; Aug 10. doi: 10.1099/jmm.0.000559.

17 Landry ML. Immunoglobulin M for Acute Infection: True or False? Clin Vaccine Immunol 2016;23:540–5. doi:10.1128/CVI.00211-16

18 Martins TB, Jaskowski TD, Mouritsen LC, Hill HR. An evaluation of the effectiveness of three immunoglobulin G (IgG) removal procedures for routine IgM serological testing. Clin Diagn Lab Immunol 1995; 2: 98–103

19 Series TS, Assessment D. Diagnostic Assessment TSS-6 Syphilis rapid diagnostic tests. Geneva World Health Organisation 2018 17 Licence CC BY-NC-SA 30 IGO 2018.

